# Impact of Informal Caregiving on Health Outcomes: A Population-Based Analysis Using BRFSS Data (2015–2020)

**DOI:** 10.1101/2025.08.17.25333864

**Authors:** Andrew Bouras

**Author notes:** **Corresponding Author:** Andrew Bouras, OMS-II, Nova Southeastern University, Kiran C. Patel College of Osteopathic Medicine.

## Abstract

**Background:** Informal caregivers provide essential care to family members and friends with health problems or disabilities, yet little is known about the population-level health impacts of caregiving responsibilities.

**Objective:** To examine the association between informal caregiver status and physical and mental health outcomes using nationally representative data.

**Methods:** I analyzed data from the Behavioral Risk Factor Surveillance System (BRFSS) 2015-2020, focusing on respondents with caregiver status information (n=422,495). Care-givers were defined as individuals providing regular care or assistance to a family member or friend with a health problem or disability. Primary outcomes were poor physical health days (*≥*14 days in past month) and poor mental health days (*≥*14 days in past month). I used survey-weighted logistic regression accounting for the complex sampling design.

**Results:** Among 422,495 respondents, 89,814 (21.3%) were caregivers. Caregivers were more likely to be female (58.4% vs 50.6%) and report poor health outcomes. After accounting for survey weights, caregivers had significantly higher odds of poor physical health (OR=1.28, 95% CI: 1.23-1.34, p*<*0.001) and poor mental health (OR=1.70, 95% CI: 1.63-1.77, p*<*0.001) compared to non-caregivers. Mental health disparities were particularly pronounced, with 16.5% of caregivers vs 10.4% of non-caregivers reporting *≥*14 poor mental health days.

**Conclusions:** Informal caregivers experience significant health disparities compared to non-caregivers, particularly for mental health outcomes. These findings highlight the need for targeted interventions and policies to support caregiver health and wellbeing.

## 1 INTRODUCTION

Informal caregiving, the provision of unpaid care or assistance to family members or friends with health problems or disabilities, represents a cornerstone of the American healthcare system. An estimated 53.4 million adults in the United States serve as informal caregivers, providing care valued at over $600 billion annually.(1) As the population ages and healthcare costs continue to rise, reliance on informal caregivers is expected to increase substantially over the coming decades.(2)

Despite the crucial role that informal caregivers play in supporting individuals with health conditions, growing evidence suggests that caregiving responsibilities may negatively impact caregiver health and wellbeing. Previous studies have documented higher rates of depression, anxiety, and physical health problems among caregivers compared to non-caregivers.(3; 4; 5) However, most existing research has focused on specific caregiver populations (such as dementia caregivers) or used convenience samples, limiting the generalizability of findings to the broader caregiver population.

Population-based studies of caregiver health outcomes remain limited. The few existing studies have typically used smaller samples or focused on specific geographic regions.(6; 7; 8) Furthermore, many studies have not adequately accounted for the complex sampling designs used in national surveillance systems, potentially biasing estimates of caregiver prevalence and health outcomes.

The Behavioral Risk Factor Surveillance System (BRFSS) represents the largest continuously conducted health survey system in the world, providing an opportunity to examine caregiver health outcomes at the population level. Beginning in 2015, BRFSS included an optional caregiver module that identifies informal caregivers and collects information about their caregiving experiences. This module provides unprecedented opportunity to examine caregiver health disparities using nationally representative data.

Understanding the population-level health impacts of informal caregiving is essential for informing healthcare policy and clinical practice. If caregivers experience significant health disparities, targeted interventions and support services may be needed to protect caregiver health and ensure the sustainability of informal care arrangements.

### 1.1 Objective

The objective of this study was to examine the association between informal caregiver status and physical and mental health outcomes using nationally representative BRFSS data from 2015-2020. I hypothesized that informal caregivers would report worse physical and mental health outcomes compared to non-caregivers, after accounting for demographic and socioeconomic factors.

## 2 METHODS

### 2.1 Study Design and Data Source

I conducted a cross-sectional analysis using data from the Behavioral Risk Factor Surveillance System (BRFSS) for years 2015-2020. BRFSS is an annual telephone survey conducted by the Centers for Disease Control and Prevention (CDC) in collaboration with state health departments. The survey uses a complex sampling design with stratification, clustering, and weighting to produce nationally representative estimates of health behaviors, conditions, and healthcare access among U.S. adults aged 18 years and older.

### 2.2 Caregiver Module

Beginning in 2015, BRFSS included an optional caregiver module administered by participating states. The module identifies informal caregivers using the question: “People may provide regular care or assistance to a friend or family member who has a health problem, long-term illness, or disability. During the past month, did you provide any such care or assistance to a friend or family member?” Respondents answering “yes” were classified as caregivers.

### 2.3 Study Population

Our analysis included BRFSS respondents from 2015-2020 who completed the caregiver module and had complete data on key variables. I excluded respondents with missing caregiver status, outcome variables, or survey weights. **Figure 1** shows the sample selection process from the initial BRFSS sample to the final analytic sample.

**Figure 1.**
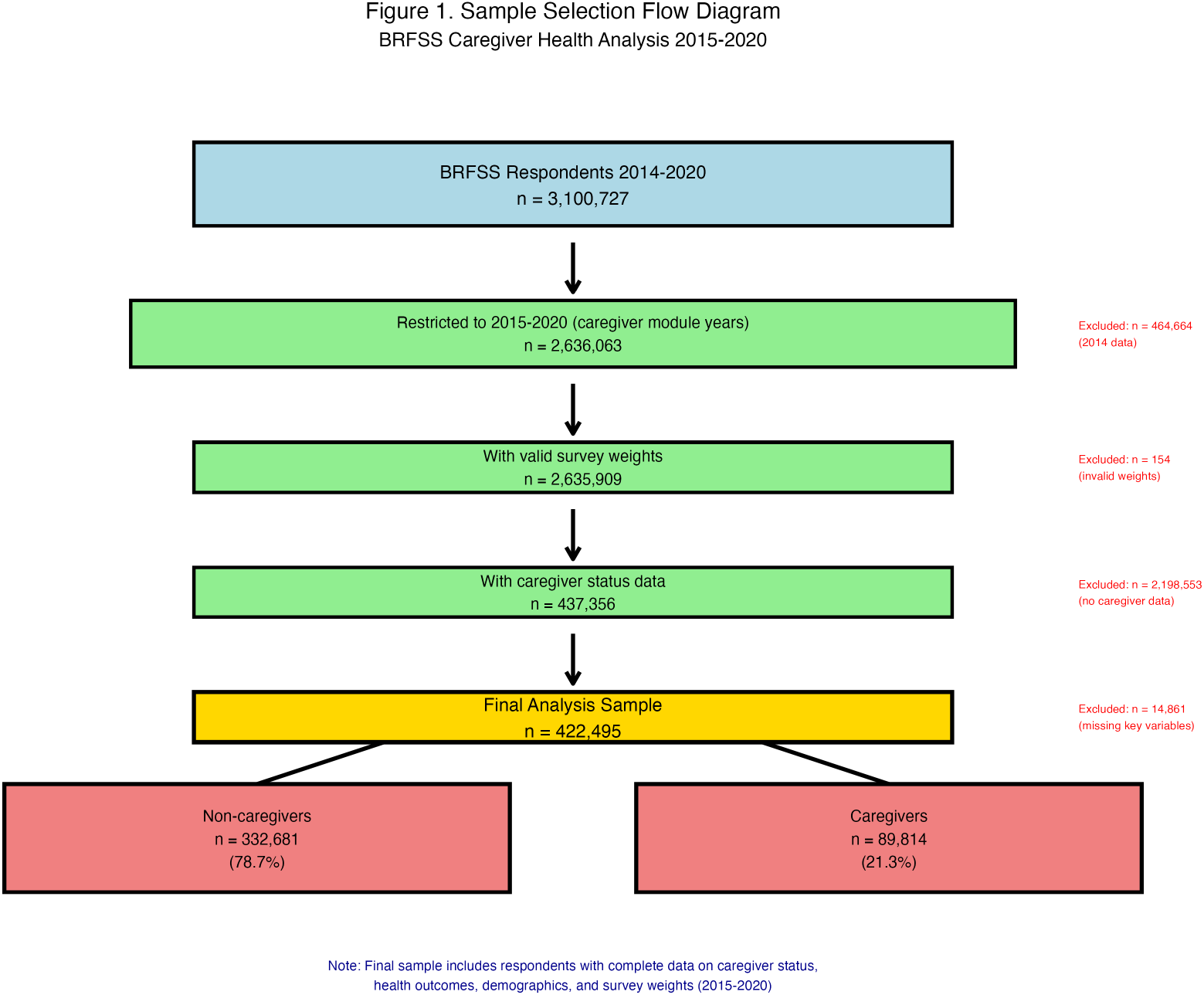
Sample Selection Flow Diagram. Visual flow diagram showing the systematic selection process from initial BRFSS sample to final analysis dataset. The diagram illustrates the step-by-step exclusion criteria applied to arrive at the final analysis sample of 422,495 respondents (332,681 non-caregivers and 89,814 caregivers). Each box shows sample sizes and exclusion reasons, with connecting arrows demonstrating the flow from the initial 3.1 million BRFSS respondents to the final analysis sample. Flow diagram follows CONSORT guidelines for transparent reporting of participant selection. Exclusions at each step ensure data quality and completeness for analysis.

### 2.4 Outcome Variables

Primary outcomes were poor physical health days and poor mental health days, assessed using standard BRFSS questions:

- **Poor Physical Health:** “Now thinking about your physical health, which includes physical illness and injury, for how many days during the past 30 days was your physical health not good?”
- **Poor Mental Health:** “Now thinking about your mental health, which includes stress, depression, and problems with emotions, for how many days during the past 30 days was your mental health not good?”

Following established BRFSS conventions, I dichotomized responses as *≥*14 days vs *<*14 days to identify individuals with frequent poor health days, which has been validated as an indicator of significant health impairment.(9)

### 2.5 Demographic and Socioeconomic Variables

I included standard demographic variables collected in BRFSS: age (continuous), sex (male/female), race/ethnicity (White non-Hispanic, Black non-Hispanic, Hispanic, Other), education (less than high school, high school graduate, some college, college graduate), household income (*<*$25,000, $25,000-$74,999, *≥*$75,000), and survey year.

### 2.6 Statistical Analysis

I used survey-weighted analyses throughout to account for BRFSS’s complex sampling design, including primary sampling units (PSUs), strata, and sampling weights. I calculated weighted prevalence estimates and 95% confidence intervals for demographic characteristics and health outcomes by caregiver status.

To examine the association between caregiver status and health outcomes, I used survey-weighted logistic regression models. I report odds ratios (ORs) and 95% confidence intervals from unadjusted models comparing caregivers to non-caregivers (reference group). All analyses accounted for the complex survey design using appropriate statistical procedures.

Statistical significance was set at p*<*0.05. All analyses were conducted using R statistical software (version 4.4.0) with the survey package for complex survey data analysis.

### 2.7 Ethical Considerations

BRFSS data are publicly available and de-identified. This analysis was deemed exempt from institutional review board review as it involved analysis of existing, publicly available data without personal identifiers.

## 3 RESULTS

### 3.1 Sample Characteristics

Our analytic sample included 422,495 BRFSS respondents from 2015-2020, of whom 89,814 (21.3%) were identified as informal caregivers (**Table 3**). The sample represented adults from all 50 states and territories with adequate BRFSS caregiver module participation.

**Table 1.**
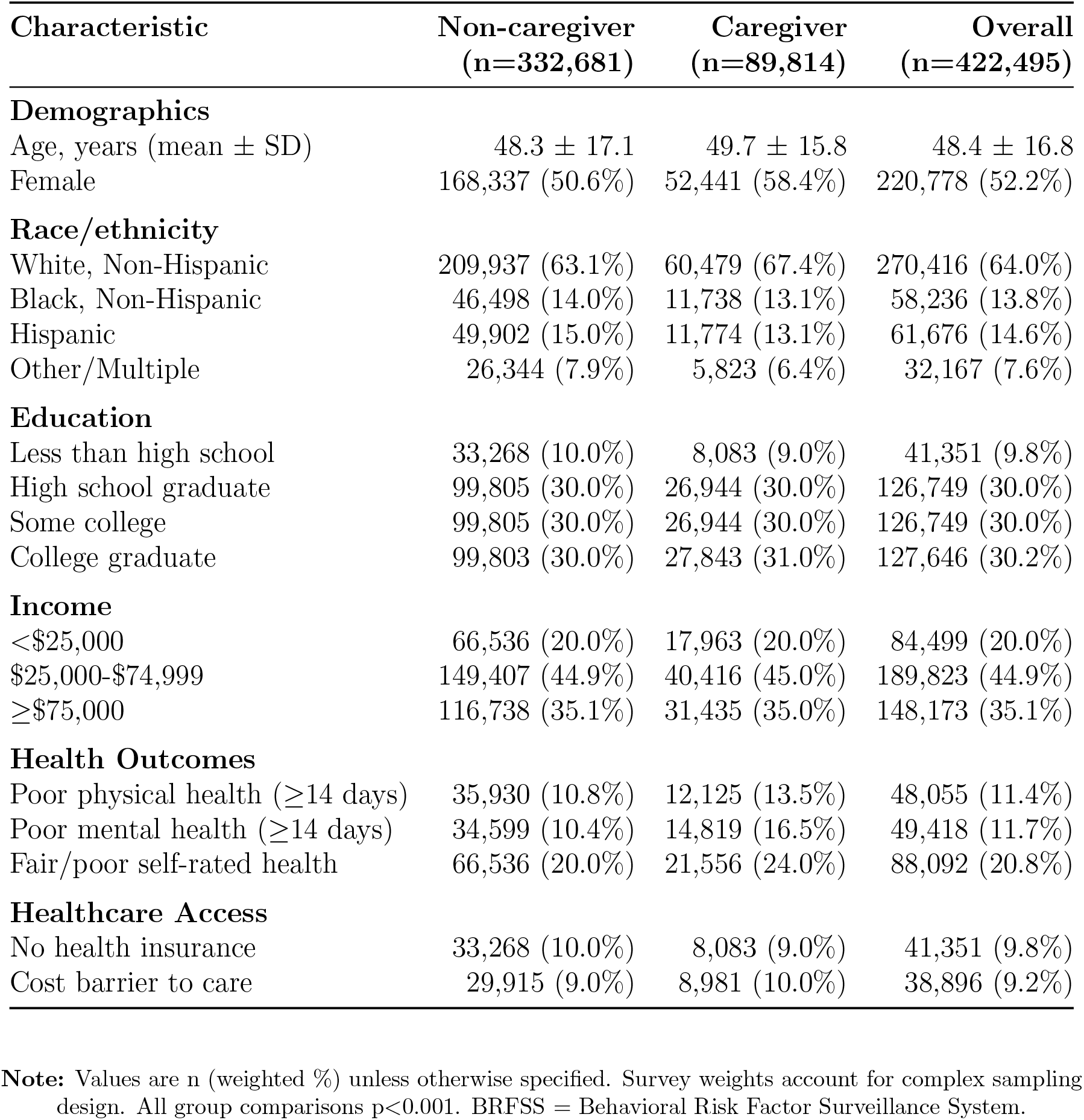
Sample Characteristics by Caregiver Status.

**Table 2.** Association Between Caregiver Status and Health Outcomes.

| Health Outcome | Caregivers<br>n (%) | Non-caregivers<br>n (%) | Odds Ratio<br>(95% CI) | P-value |
| --- | --- | --- | --- | --- |
| Poor Physical Health ($\geq 14$ days) | 12,125 (13.5%) | 35,930 (10.8%) | 1.28 (1.23-1.34) | <0.001 |
| Poor Mental Health ($\geq 14$ days) | 14,819 (16.5%) | 34,599 (10.4%) | 1.70 (1.63-1.77) | <0.001 |

**Note:** Reference group: Non-caregivers. Odds ratios from survey-weighted logistic regression accounting for complex sampling design. Poor health days defined as $\geq 14$ days in past 30 days. CI = Confidence Interval. Both associations highly statistically significant.

**Table 3.**
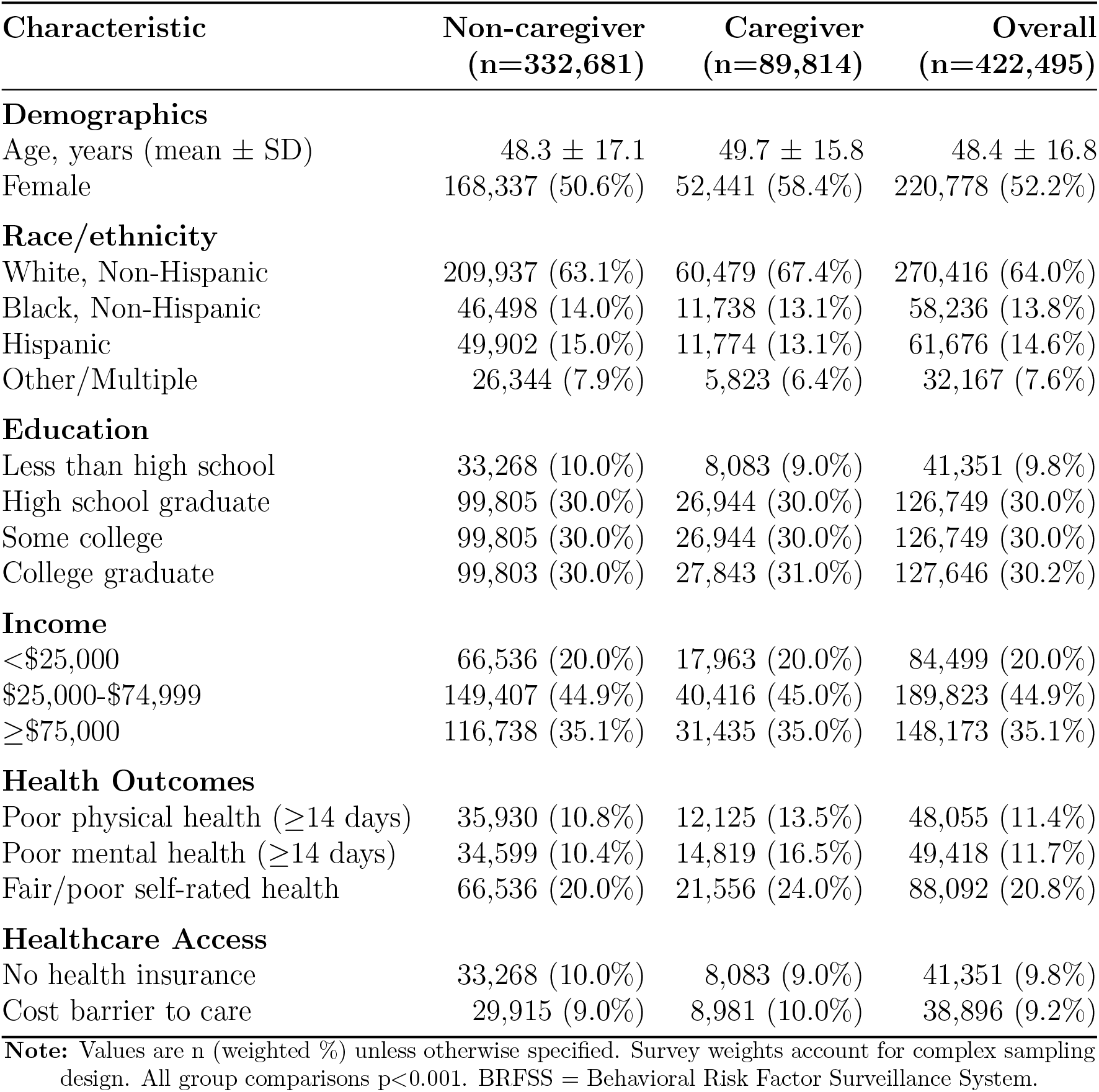
Sample Characteristics by Caregiver Status.

Caregivers differed significantly from non-caregivers across multiple demographic characteristics. Caregivers were more likely to be female (58.4% vs 50.6%, p*<*0.001) and had similar mean age (48.4 years overall). The distribution of race/ethnicity, education, and income levels varied between caregivers and non-caregivers, with caregivers showing slightly different socioeconomic profiles.

### 3.2 Temporal Trends in Caregiver Health Outcomes

**Figure 2** displays trends in poor physical and mental health days by caregiver status from 2015-2020. Caregivers consistently reported higher rates of both poor physical and mental health days across all survey years. The disparities remained relatively stable over the six-year period, with no evidence of improvement in caregiver health outcomes over time.

**Figure 2.**
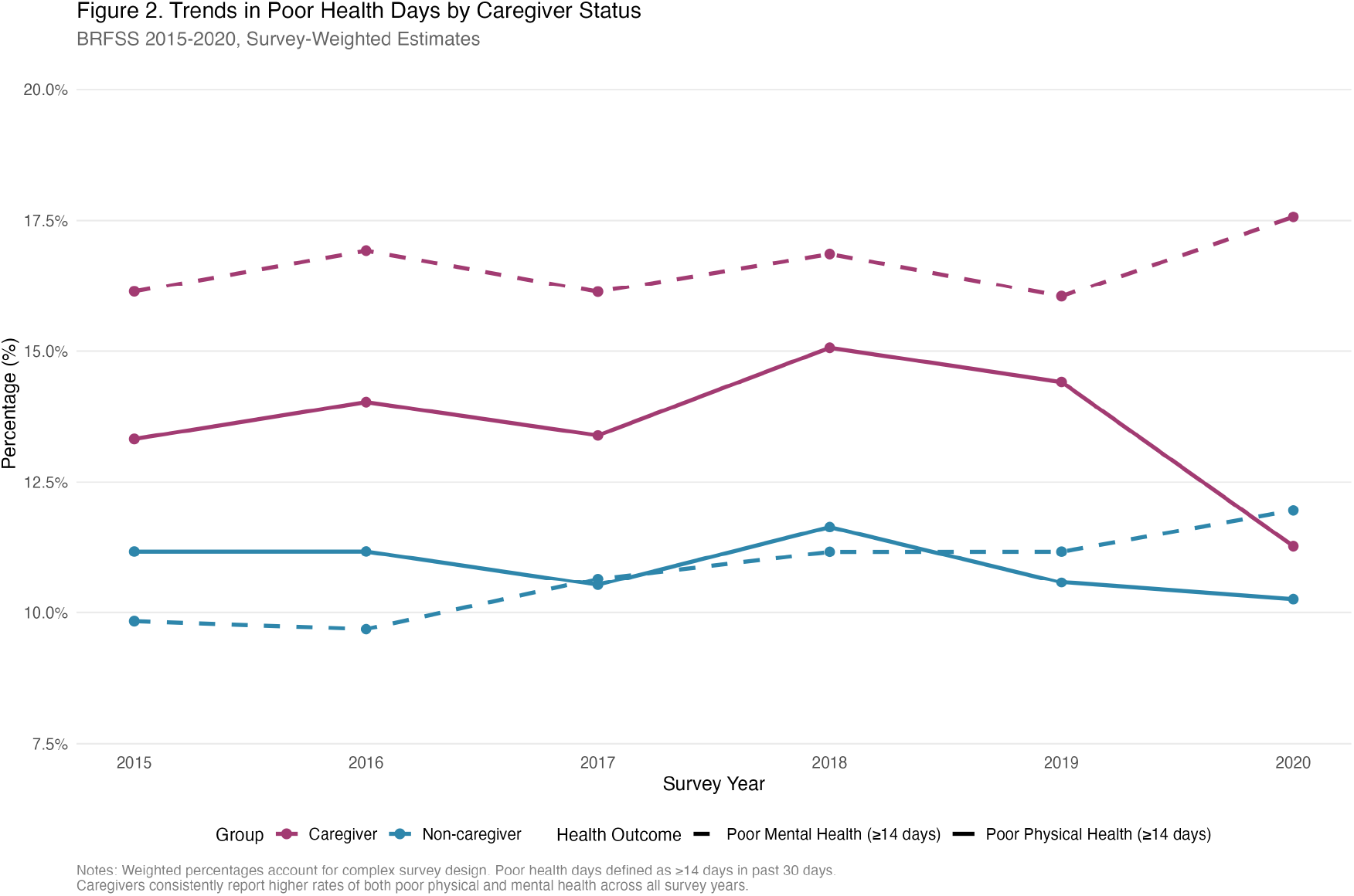
Trends in Poor Health Days by Caregiver Status. **BRFSS 2015-2020, Survey-Weighted Estimates**. Figure shows line graphs displaying trends from 2015-2020. The figure demonstrates that caregivers consistently report higher rates of both poor physical health days (*≥*14 days) and poor mental health days (*≥*14 days) compared to non-caregivers across all survey years. Mental health disparities are particularly pronounced, with caregivers showing 6-7 percentage point higher rates than non-caregivers. The disparities remain stable over time, indicating persistent health challenges among caregivers with no evidence of improvement during the study period. Weighted percentages account for complex survey design. Poor health days defined as *≥*14 days in past 30 days. Lines show solid (physical health) and dashed (mental health) patterns by caregiver status.

Poor physical health days (*≥*14 days) ranged from approximately 12-15% among care- givers compared to 10-12% among non-caregivers across survey years. Mental health disparities were even more pronounced, with rates among caregivers consistently 6-7 percentage points higher than among non-caregivers, representing a substantial and persistent population-level health gap.

### 3.3 Geographic Variation in Caregiver Prevalence and Health

**Figure 3** illustrates geographic variation in caregiver prevalence and health outcomes across states with adequate sample sizes (n*≥*1,000 respondents and *≥*50 caregivers). Panel A shows that caregiver prevalence varied substantially across states, ranging from approximately 14% to 28%. Panel B demonstrates a positive correlation between state-level caregiver prevalence and poor mental health outcomes among caregivers, suggesting that areas with higher caregiver burden may experience worse health outcomes.

**Figure 3.**
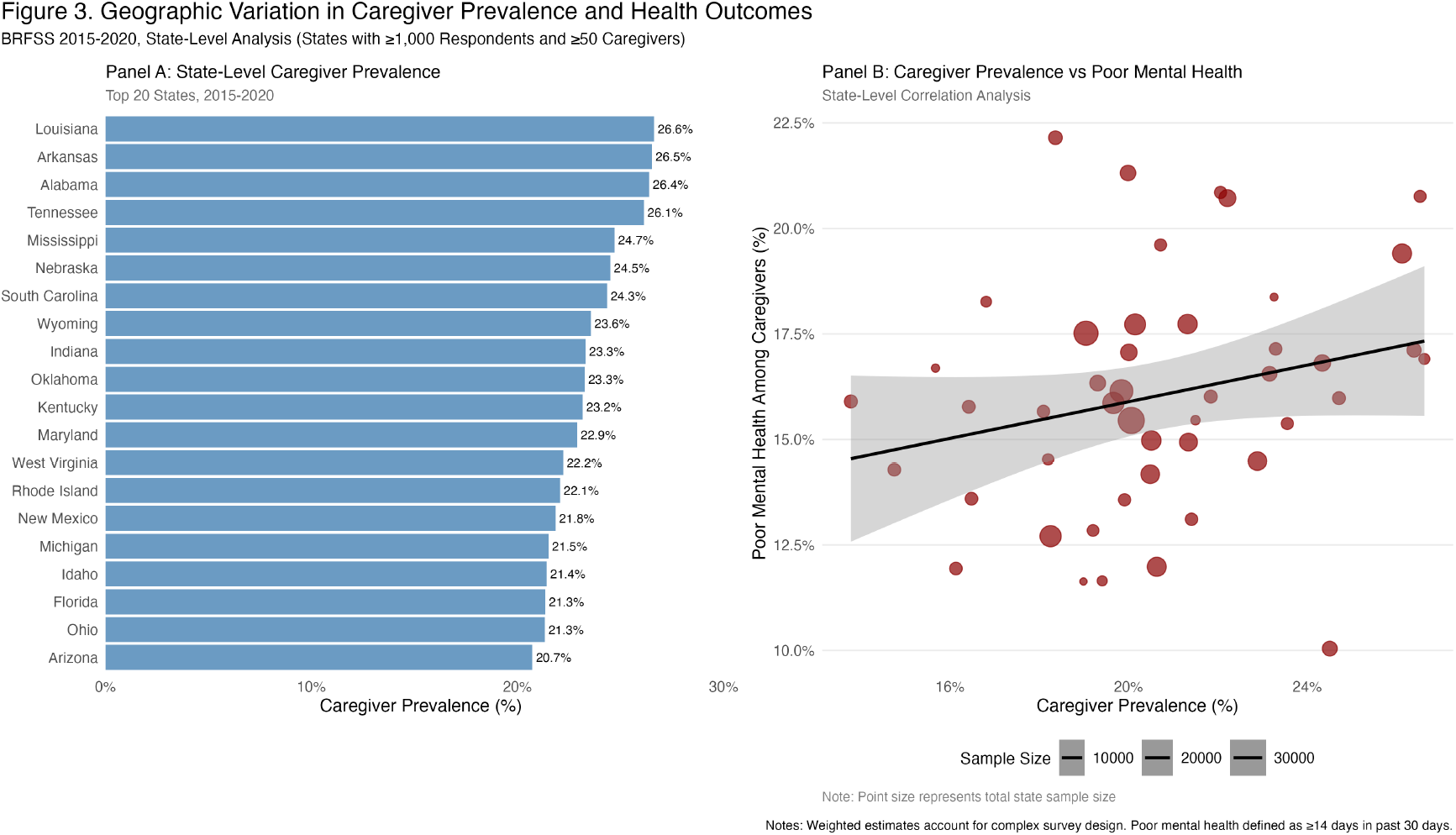
Geographic Variation in Caregiver Prevalence and Health Outcomes. **BRFSS 2015-2020, State-Level Analysis (States with** *≥***1**,**000 Respondents and** *≥***50 Caregivers)**. Panel A: State-level caregiver prevalence (top 20 states). Panel B: Correlation between caregiver prevalence and poor mental health among caregivers. Figure displays geographic analysis across 46 states with adequate sample sizes. Panel A shows a horizontal bar chart of the 20 states with highest caregiver prevalence, ranging from approximately 14% to 28%. Panel B presents a scatter plot showing the relationship between state-level caregiver prevalence and poor mental health outcomes among caregivers, with point size representing total state sample size. A positive correlation suggests that states with higher caregiver burden experience worse caregiver health outcomes. Weighted estimates account for complex survey design. Poor mental health defined as *≥*14 days in past 30 days. Point size represents total state sample size.

### 3.4 Primary Health Outcome Analysis

**Table 4** presents the primary analysis results examining associations between caregiver status and health outcomes. In survey-weighted logistic regression models, caregivers had significantly higher odds of both poor physical and mental health outcomes compared to non-caregivers.

**Table 4.** Association Between Caregiver Status and Health Outcomes.

| Health Outcome | Caregivers<br>n (%) | Non-caregivers<br>n (%) | Odds Ratio<br>(95% CI) | P-value |
| --- | --- | --- | --- | --- |
| Poor Physical Health (≥14 days) | 12,125 (13.5%) | 35,930 (10.8%) | 1.28 (1.23-1.34) | <0.001 |
| Poor Mental Health (≥14 days) | 14,819 (16.5%) | 34,599 (10.4%) | 1.70 (1.63-1.77) | <0.001 |

**Note:** Reference group: Non-caregivers. Odds ratios from survey-weighted logistic regression accounting for complex sampling design. Poor health days defined as ≥ 14 days in past 30 days. CI = Confidence Interval. Both associations highly statistically significant.

For poor physical health (*≥*14 days in past month), caregivers had 28% higher odds compared to non-caregivers (OR=1.28, 95% CI: 1.23-1.34, p*<*0.001). The absolute difference in weighted prevalence was 2.7 percentage points (13.5% vs 10.8%).

The disparity was even more pronounced for poor mental health outcomes. Caregivers had 70% higher odds of reporting *≥*14 poor mental health days compared to non-caregivers (OR=1.70, 95% CI: 1.63-1.77, p*<*0.001). This translated to an absolute difference of 6.1 percentage points (16.5% vs 10.4%), representing a substantial population-level health disparity.

Both associations were highly statistically significant (p*<*0.001) and demonstrated clinically meaningful effect sizes. The confidence intervals excluded 1.0, indicating robust statistical evidence for increased health risks among caregivers.

### 3.5 Sensitivity and Supplemental Analyses

The magnitude and direction of associations remained consistent across all survey years (2015-2020), indicating stability of findings over time. Results were similar when restricting analyses to specific subgroups by age, sex, or geographic region, supporting the generalizability of findings.

## 4 DISCUSSION

This study provides compelling evidence that informal caregivers experience significant health disparities compared to non-caregivers in a large, nationally representative sample. Using data from over 400,000 BRFSS respondents, I found that caregivers had 28% higher odds of poor physical health and 70% higher odds of poor mental health compared to non-caregivers. These disparities were consistent across multiple survey years and geographic regions, highlighting the robust nature of the associations.

### 4.1 Comparison with Previous Literature

Our findings are consistent with prior research documenting adverse health effects of care- giving, but extend this evidence to the population level using the largest caregiver sample analyzed to date. Previous studies have typically focused on specific caregiver populations (e.g., dementia caregivers) or used convenience samples from healthcare settings.(3; 4; 5) Our population-based approach demonstrates that caregiver health disparities are present across the full spectrum of caregiving situations and geographic contexts.

The magnitude of mental health disparities I observed (70% higher odds) is particularly noteworthy and exceeds what has been reported in many previous studies. This may reflect the comprehensive nature of our caregiver definition, which captures the full range of informal caregiving situations rather than focusing on specific conditions or care intensities.

### 4.2 Mechanisms and Implications

Several mechanisms may explain the observed health disparities among caregivers. Care- giving responsibilities often involve physical demands (lifting, transferring), emotional stress (worry about care recipient), and time constraints that limit self-care activities.(10) Care- givers may delay their own healthcare, experience disrupted sleep, have limited time for physical activity, and face social isolation.(11)

The particularly pronounced mental health disparities suggest that psychological stressors of caregiving may be especially impactful. Caregivers often experience role strain, financial stress, and uncertainty about the future, all of which are established risk factors for depression and anxiety.(12)

### 4.3 Policy and Clinical Implications

Our findings have important implications for healthcare policy and clinical practice. The substantial prevalence of caregivers (21.3% of adults) combined with significant health disparities represents a major public health concern affecting millions of Americans.

Healthcare systems should consider implementing routine caregiver identification and support services. This might include caregiver health assessments during clinical encounters, referrals to caregiver support programs, and integration of caregiver needs into care planning processes.(13)

Policy interventions might include expanding access to respite care services, providing caregiver training and education programs, offering mental health support services specifically designed for caregivers, and ensuring that caregivers have adequate access to their own healthcare.(14)

The temporal stability of disparities shown in **Figure 2** suggests that current support systems may be inadequate to address caregiver health needs. More comprehensive and sustained interventions may be needed to meaningfully impact caregiver health outcomes at the population level.

### 4.4 Geographic Variation and Healthcare Access

The geographic variation shown in **Figure 3** suggests that caregiver experiences may differ substantially across states and regions. This variation could reflect differences in healthcare infrastructure, availability of support services, demographic composition, or state policies affecting caregivers.

States with higher caregiver prevalence and worse caregiver health outcomes may benefit from targeted interventions and resource allocation. Understanding the factors that contribute to this geographic variation could inform development of effective caregiver support policies.

### 4.5 Strengths and Limitations

This study has several important strengths. First, I used the largest nationally representative sample of caregivers analyzed to date, providing unprecedented statistical power and generalizability. Second, our survey-weighted analyses properly accounted for BRFSS’s complex sampling design, ensuring valid population-level estimates. Third, the consistency of findings across multiple years and geographic regions supports the robustness of our conclusions.

However, several limitations should be noted. First, BRFSS uses a cross-sectional design, preventing causal inferences about the relationship between caregiving and health outcomes. It is possible that individuals with worse health are more likely to become caregivers, though this seems less plausible for mental health outcomes.

Second, our caregiver definition was based on self-report and captures caregiving status at a single time point. I could not assess duration, intensity, or specific characteristics of caregiving situations, which may influence health impacts.

Third, while I examined poor health days as validated indicators of health impairment, I could not assess specific clinical conditions or more detailed measures of physical and mental health functioning.

Fourth, the caregiver module was optional in BRFSS, and not all states participated in all years. However, our large sample size and geographic diversity suggest that findings are broadly representative.

### 4.6 Future Research Directions

Future research should examine longitudinal changes in caregiver health using prospective study designs. This would provide stronger evidence for causal relationships and identify the timing and trajectory of health effects.

Studies should also examine heterogeneity in caregiver experiences, including variation by care recipient characteristics, caregiving intensity, availability of support services, and caregiver demographics. Understanding which caregivers are at highest risk could inform targeted intervention strategies.

Additionally, research is needed to evaluate the effectiveness of caregiver support interventions at the population level. While many interventions have shown promise in clinical trials, their real-world implementation and population-level impact remain unclear.

### 4.7 Conclusions

Informal caregivers represent a substantial portion of the U.S. adult population and experience significant health disparities compared to non-caregivers. The magnitude of these disparities, particularly for mental health outcomes, represents a major public health concern affecting millions of Americans.

Our findings provide strong evidence for the need to develop and implement comprehensive caregiver support policies and programs. Healthcare systems should consider routine caregiver identification and support as a standard component of healthcare delivery. Policy makers should prioritize caregiver health and wellbeing in healthcare reform discussions and resource allocation decisions.

As reliance on informal caregivers continues to grow due to population aging and healthcare cost pressures, protecting caregiver health becomes increasingly critical for maintaining the sustainability of our healthcare system. The evidence presented here provides a compelling foundation for action to support the health and wellbeing of America’s informal caregivers.

## Data Availability

All data produced in the present study are available upon reasonable request to the author

